# Endonuclease-based genotyping of the RBM, a first-line method for the surveillance of emergence or evolution of SARS-CoV-2 Variants

**DOI:** 10.1101/2021.06.16.21259035

**Authors:** Eva Lopez, Margot Barthélémy, Cécile Baronti, Shirley Masse, Alessandra Falchi, Fabien Durbesson, Renaud Vincentelli, Xavier de Lamballerie, Rémi Charrel, Bruno Coutard

## Abstract

Since the beginning of the Covid-19 pandemics, variants have emerged. Whereas most of them have no to limited selective advantage, some display increased transmissibility and/or resistance to immune response. To date, most of the mutations involved in the functional adaptation are found in the Receptor Binding Module (RBM), close to the interface with the human receptor ACE2. In this study, we thus developed and validated a fast and simple molecular assay allowing the detection and partial identification of the mutations in the RBM coding sequence. After the amplification of the region of interest, the amplicon is heat-denatured and hybridized with an amplicon of reference. The presence of a mutation in the heteroduplex can be cleaved by a mismatch-specific endonuclease and the cleavage pattern is analysed by capillary electrophoresis. The approach was first validated on viral RNA purified different SARS-CoV-2 variants produced in the lab before being implemented for clinical samples. The results highlighted the performance of the assay for the detection of mutations in the RBM from clinical samples. The procedure can be easily set up for high throughput identification of the presence of mutations and serve as a first-line screening to select the samples for full genome sequencing.

## Introduction

In December 2019, individuals with pneumonia of unknown aetiology were recorded in the city of Wuhan, China. The number of cases increased steadily in the following weeks including in the countries surrounding China (Taiwan, Thailand, Malaysia, etc.), and then throughout the world via aerial transport, triggering the WHO to announce a Public Health Emergency of International Concern (PHEIC) on the 1^st^ of February 2020. Shortly after, the disease was named “COVID-19”, for “Coronavirus disease 2019”. In April 2021, more than 176 million people contracted the virus and 3.8 million died. The coronavirus in question, SARS-CoV-2, is an enveloped, positive single stranded RNA virus. The viral particle exposes at its surface the envelope glycoprotein Spike (S). The S protein is a multi-domain protein (Figure 1A) involved in host cell recognition, in particular *via* its receptor binding domain (RBD) which specifically binds the human Angiotensin-2 Converting Enzyme (hACE2). The S protein is considered a major determinant of viral infectivity and antigenicity [1] [2], and mutations in the coding sequence of the S protein are susceptible to affect the biology of SARS-CoV-2.

**Figure 1:**
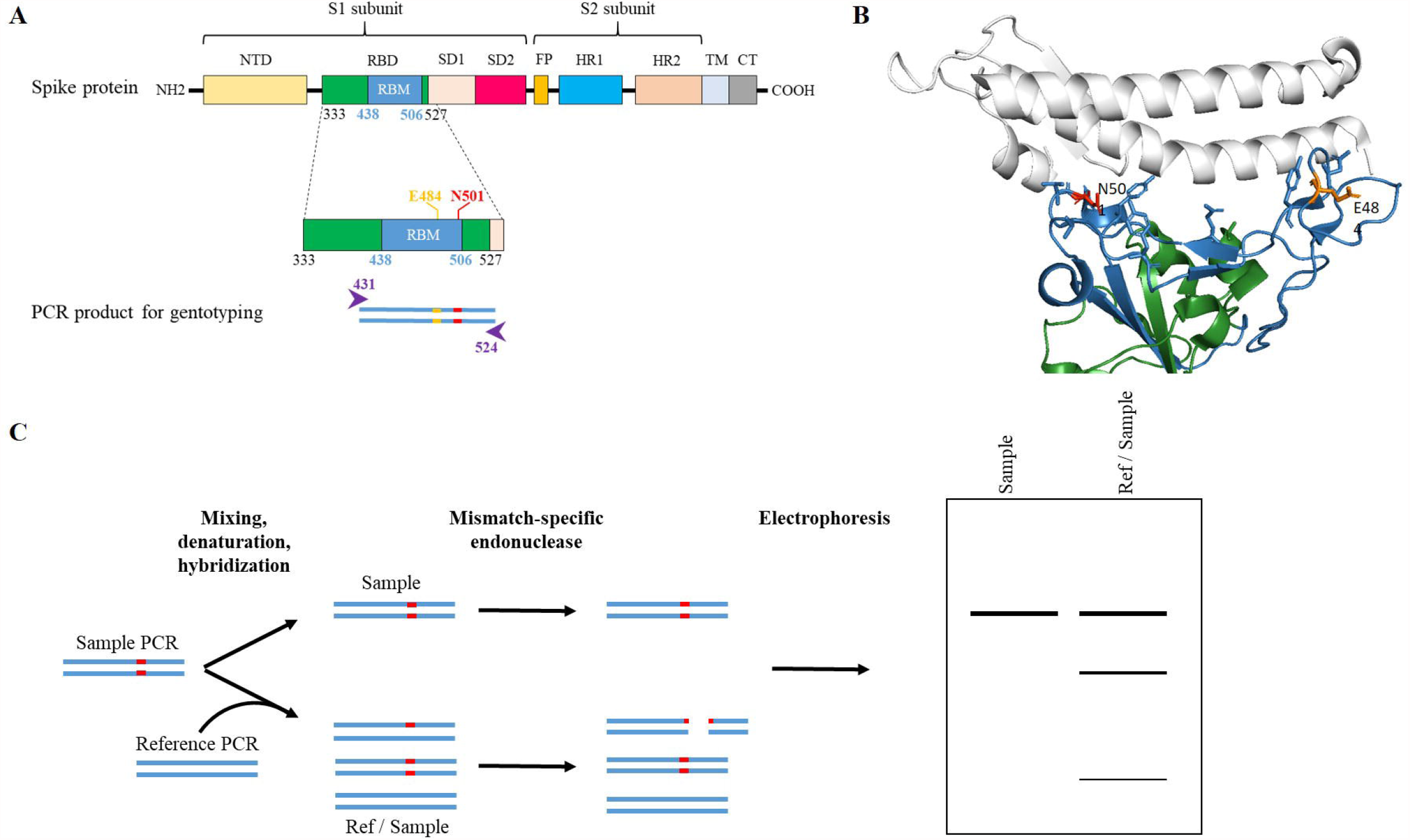
RBM is a key region for viral for viral entry and seroneutralization. A: Primary structure of the SARS-CoV-2 Spike. NTD: N-terminal domain; RBD: Receptor binding domain; RBM: Receptor binding motif; SD1: Subdomain 1; SD2: Subdomain 2; FP: Fusion peptide; HR1: Heptad repeat 1; HR2: Heptad repeat 2; TM: Transmembrane region; CT: Cytoplasmic region. Amino acid positions of the RBD and RBM boundaries are presented in black and blue numbers, respectively. Functional mutations in the RBM are shown in orange (E484K) and red (N501Y). Purple arrows and numbers correspond to the RT-PCR amplified region. B: Structure representation of the interaction between SARS-CoV-2 RBD core (green) and hACE2 receptor (white) (PDB 6vw1). RBM is shown in blue. E484 and N501 positions are highlighted in orange and red, respectively. C: Detection of mutation(s) in a sample compared to a reference sequence. Results of the digestion by a mismatch-specific endonuclease of an auto-control (amplicon of a sample alone) and mix (amplicons of reference and sample) are analysed by electrophoresis and fragment patterns are detected.

Since the emergence of SARS-CoV-2 several non-synonymous mutations with biological implications have been reported in the coding sequence of the S protein. For instance the first one led to D614G, a substitution contributing to the enhancement of viral loads in the upper respiratory tract with possible increased transmission [3]. Latter, several variants designed as Variant of Concern (VoC) have emerged and are disseminating. Those variants may demonstrate increased transmissibility or severity of the disease, reduction of sero-neutralization by antibodies induced by previous infection or vaccination, or resistance to therapeutic treatments. Among the VoCs, the first one - 501Y.V1 or B.1.1.7 lineage - was identified in the UK and showed enhanced human-to-human transmission and increased disease severity [4] [5]. Then, variants of B1.351 (501Y.V2), P.1 (501Y.V3) and B.617 lineages were isolated and characterized in South Africa, Brazil/Japan and India, respectively. Both B1.351 (501Y.V2), P.1 (501Y.V3) variants show increased resistance to antibody neutralization [5]–[7]. VoCs have in common to present at least one non-synonymous mutation in the spike receptor binding motif (RBM). RBM is the sub-domain of the RBD containing most of the hACE2-contacting residues and is also characterized by the presence of epitopes for neutralising antibodies (Figure 1B). Some of the characterized mutations are N501Y for 501Y.V1, E484K/N501Y for 501Y.V2 and 501Y.V3. By themselves, these mutations can affect the binding of the S protein to hACE2 and/or the potency of neutralizing antibodies [8]–[12]. The genetic evolution of this region is specifically scrutinized to identify possible new VoCs. The rapid detection of VoCs is thus pivotal for mitigating transmission in hospital settings and for adjusting therapies to avoid lowering efficacy.

The detection of VoCs and surveillance of the evolution of SARS-CoV-2 population are currently surveyed by two approaches: variant-specific real time RT-PCR for the search of mutations at given positions and massive campaigns of New Generation Sequencing (NGS), whose data can contribute to the public-health decision making [13]. On one side, real time RT-PCR is fast and operational on site, but it can detect only known mutations and does not address the newly emerging ones. On the other side, NGS can detect any mutation along the genome but the results is obtained in days rather than hours, delaying information required for medical decision to be taken upon sequence identification. In addition, all the biological samples cannot be sequenced and upstream sampling is mandatory for the selection of the most relevant biological samples to characterize. Here we present the proof-of-concept for an alternative method allowing the surveillance of the genetic drift of SARS-CoV-2 in the RBM region where mutations are susceptible to affect the dissemination, pathogenicity or antibody-resistance of the virus. The technique, relying on the amplification of the RBM coding sequence followed by an assay using a mismatch-specific endonuclease, has been validated on biological samples demonstrating its feasibility.

## Material and methods

### Choice of the target sequence for amplification

The target of the SARS-CoV-2 Spike coding sequence ranges from nt position 1273 to 1587, including the primers, and is 315 nucleotide long. The corresponding amino acid sequence (amino acid 425 to 529) encompasses the RBM module (Figure 1).

### Purified viral RNA from infected cell cultures

VeroE6/TMPRSS2^+^ (CFAR#100978) cells were grown in minimal essential medium (MEM) (Life Technologies) with 7□.5% heat-inactivated foetal calf serum (FCS; Life Technologies), at 37°C with 5% CO_2_ with 1% penicillin/streptomycin (PS, 5000 U.mL^−1^ and 5000 µg.mL^−1^ respectively; Life Technologies), supplemented with 1□% non-essential amino acids (Life Technologies) and L-Glutamine (Life Technologies). SARS-CoV-2 strain BavPat1 was obtained from Pr. C. Drosten through EVA GLOBAL (https://www.european-virus-archive.com/). SARS-CoV-2 strain 2021/FR/7b-ex UK (EVA-G ref: 001V-04044) belonging to lineage 501Y.V1 and strain 2021/FR/1299-ex SA (EVA-G ref: 001V-04067) belonging to 501Y.V2 were isolated from human nasopharyngeal swabs. The virus stock was produced on VeroE6/TMPRSS2+ cells. Briefly, a 25 cm^2^ flask of subconfluent cells was inoculated with each strain at MOI=0.001. Cells were incubated at 37 °C overnight, after which the medium was changed, and incubation was continued for 24 h. Supernatant was collected, clarified by spinning at 1500× g for 10 min, supplemented with 25mM HEPES (Sigma-Aldrich) and stored at −80°C in several aliquots. All infectious experiments were conducted in a biosafety level (BSL) 3 laboratory. RNA extraction was performed using the QIAamp 96 DNA kit and the Qiacube HT plasticware kit on the Qiacube HT automate (Qiagen). Viral RNA obtained was quantified by real-time RT-PCR (EXPRESS One-Step Superscript qRT-PCR Kit (Invitrogen)) using 3.5 µL of RNA and 6.5 µL of RT qPCR mix and standard fast cycling parameters, i.e., 10 min at 50 °C, 2 min at 95 °C, and 40 amplification cycles (95 °C for 3 s followed by 30 s at 60 °C) [14]. Quantification was provided by four 2 log serial dilutions of an appropriate T7-generated synthetic RNA standard of known quantities (10^2^ to 10^8^ copies). Real-time RT-PCR reactions were performed on QuantStudio 12K Flex Real-Time PCR System (APPLIED BIOSYSTEMS, Waltham, USA) and analysed using QuantStudio 12 K Flex Applied Biosystems software v1.2.3. Primers and probe sequences, which target SARS-CoV-2N gene, were: Forward: 5’-GGCCGCAAATTGCACAAT-3’; Reverse: 5’-CCAATGCGCGACATTCC-3’; Probe: FAM-CCCCCAGCGCTTCAGCGTTCT-BHQ1.

### RNA from clinical samples

For the first set of 8 clinical samples, we collected anonymised residual nasopharyngeal (NP) samples of COVID-19 patients confirmed by real time RT-PCR from clinical laboratories based in Corsica on mid-January 2021. Total nucleic acid was extracted from 200 μL of NP samples using the QIACUBE processing system with the QIAamp 96 Virus QIAcube HT Kit (Qiagen) and eluted to 100 μL of total nucleic acid. Real-time RT-PCR was performed for each sample with TaqPath COVID-19 Kit™ (ThermoFisher). The TaqPath COVID-19 Kit™ is a multiplex real time RT-PCR diagnostic assay targeting three regions of the SARS-CoV-2 genome (N, S, and ORF1ab) which was approved by the French National Center of respiratory diseases for the detection of SARS-CoV-2. TaqPath RT-PCR™ assay is a useful tool enabling a rapid screening of SARS-CoV-2 as a S-gene target failure was observed in variants with a deletion at positions 69-70 (ΔHV69-70) whereas ORF1ab and N targets were correctly amplified [15].

No nominative nor sensitive data on participant people have been collected. This study falls within the scope of the French Reference Methodology MR-004 according to 2016–41 law dated 26 January 2016 on the modernization of the French health system. The ethics committee of University of Corsica Pascal Paoli (IRB UCPP 2020-01) approved this study.

The collection of 92 SARS-CoV-2 positive clinical samples used in the second evaluation phase were kindly provided by the National Reference Center for Respiratory Viral Infections (CNR Lyon, France). The SARS-CoV-2 genomes from the 92 samples were fully sequenced by the CNR for the surveillance of the variants.

### RT-PCR of the targeted sequence

#### RT-PCR protocol

RT-PCR was performed with SuperScript II One-Step Platinium Kit (#10928042, Thermo Fisher Scientific), on a Biometra T3000 thermal cycler. Primers were synthesized and provided by ThermoFisher Scientific. The forward and reverse primer sequence were 5’-TTACCAGATGATTTTACAGGC-3’ and 5’-AGACTTTTTAGGTCCACAAAC-3’ respectively. The cycling conditions were defined as follows : 50 °C for 30 min. ; 94 °C for 2 min. ; 40 cycles of 94 °C for 15 sec. ; 55 °C for 20 sec. and 68°C for 20 sec. The final elongation was performed at 72°C for 2 min.

#### Linearity of the designed system

To determine the linearity of the designed system, real time RT-PCR was performed with QuantiTect SYBR Green RT-PCR Kit (#204243, Qiagen), on a BioRad CFX96™ thermal cycler, software version 3.0 (Bio-Rad Laboratories). The cycling conditions were : 50 °C for 30 min. ; 95 °C for 15 min. ; 40 cycles of 95 °C for 15 sec. ; 50 °C for 30 sec. and 72°C for 45 sec. PCR products were then transferred to a 2% agarose gel with added SYBR™ Safe DNA Gel Stain 1X (#S33102, ThermoFisher Scientific). The determination of the fragment size is estimated using a molecular size ladder (DNA 1kb Plus #10787026, ThermoFisher Scientific).

#### Production of the reference PCR products

SARS-CoV-2 BavPat1 strain was selected as the reference for this study. A large scale production of amplicons was done for this strain, following the RT-PCR protocol defined above.

#### Production of the PCR products from biological samples

Each sample used on this study have been amplified following the RT-PCR protocol described above.

### Mismatch-specific endocnuclease assay and detection of cleavage

The presence of a mutation on the amplified target was detected using the Surveyor® Mutation Detection Kit (#706021, Integrated DNA Technologies). Two PCR products (a reference product and a sample to be analysed) were mixed in a 10µL final volume and the endonuclease mismatch specific cleavage has been proceed following the manufacturer recommendations. The mixture was then loaded on capillary electrophoresis system (Fragment Analyser 5200, Agilent or GXII, Perkin Elmer) prior to analysis of the cleavage profile. When needed for confirmation of the presence of mutations, the PCR products were sequenced using the Sanger method with the forward and reverse primers used for the RT-PCR (Genewiz).

## Results and discussion

### Principle of the method

Non-synonymous mutations occurring in the RBM coding sequence of SARS-CoV-2 are pivotal because they are susceptible to change the phenotype with possible influence on transmission pattern, increased pathogenesis or immune escape; the latter can result in iterative infections, reduced vaccine efficacy or resistance to Mab-based therapeutics. Accordingly, RBM genetic evolution has to be monitored for the surveillance of emerging variants. To detect mutations in the RBM region, we evaluated the SURVEYOR® Nuclease S, an endonuclease cleaving double strand DNA where mismatches exist. The enzyme had already been used for the detection of mutations in *brca1* and *brca2* genes in the case of hereditary breast cancer [16]. The principle of the technique relies on the creation of heteroduplexes between an amplified DNA from a reference nucleic acid (*Ref*) and an amplified DNA from a sample to be evaluated (*Sample*). Both amplicons are mixed (Figure 1C). After denaturation/hybridization, a mixture of homoduplexes (*Ref*/*Ref* and *Sample*/*Sample*) and heteroduplexes (*Ref*/*Sample*) are produced. If the sequence from the S*ample* has mutations compared to the *Ref* sequence, mismatches happen in the heteroduplex. The latter is cleaved, resulting in cleavage products that can be evidenced by capillary electrophoresis, providing information on the number of Single Nucleotide Polymorphism (SNPs) and their approximate location(s). In contrast, if the sequences of *Ref* and *Sample* are identical no cleavage is observed and only the full-length PCR products are visible by electrophoresis.

### Sensitivity of the RT-PCR assay targeting the RBM coding sequence

Within the RBD region of the Spike protein, the RBM contains amino-acids subjected to mutations (AA) positions – *eg* 452, 484 and 501 - with functional relevance and observed in VoCs (Figure 1A). Its coding sequence was thus well-suited for a genotyping assay. Prerequisite was also that the targeted region is centred on the positions of interest and short enough so that it excludes the identification of mutations with low to no functional effect, *ie* synonymous or non-synonymous with functional consequence. The size of the PCR product was thus set to 315-nt, allowing the detection of mutations in the region of the Spike protein spanning AA positions 431 to 524 of the S protein.

To set up the assay, we first evaluated the sensitivity of the RT-PCR system in the range 1-10^6^ of RNA copies/µL. The evaluation was done either by real-time RT-PCR with SYBR green (Figure 2A, 2B), or by end-point analysis of the PCR products on agarose gel electrophoresis (Figure 2C). The amplification is linear from 10 to 10^5^ copies of RNA/µL, with correlation coefficient R^2^=0.9986 (Figure 2A), and has a limit of detection on agarose gel up to 10 copies/µL (Figure 2C). However, given the quantity of material needed for the nuclease assay (>25 ng/µL), we arbitrarily applied the threshold at 10^3^/10^4^ copies/µL, which corresponds to samples with Ct values between 28 to 30 in reference detection systems [17]. It should be noted that the SYBR green inhibits the endonuclease used for the detection of mismatches, likely by altering the structure of the DNA helix, thus rendering the resulting PCR product not suitable for subsequent capillary electrophoresis analysis (data not shown).

**Figure 2:**
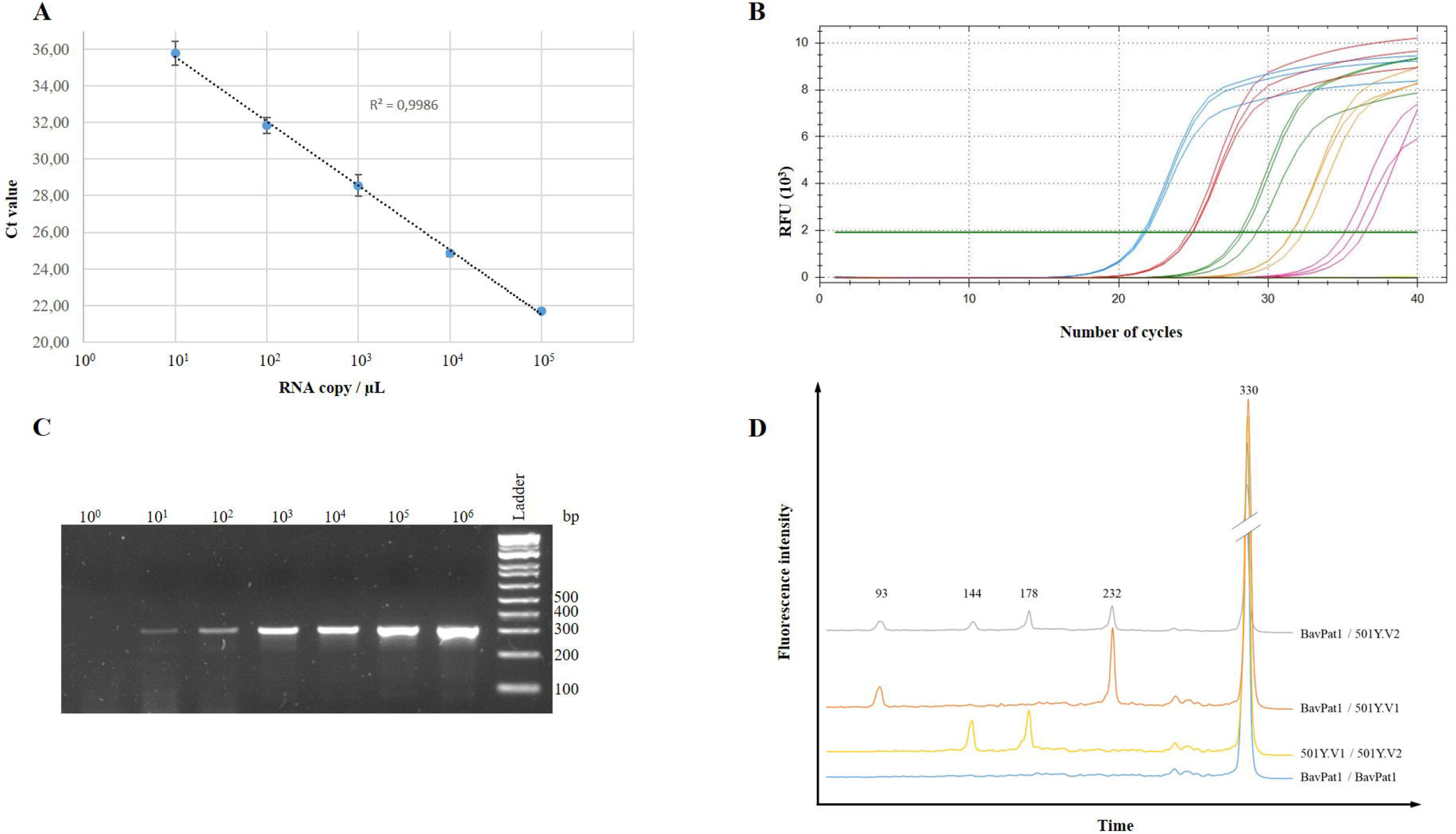
Evaluation of the target amplification and mismatch-specific nuclease assay on reference material. A and B: Linearity of the real time RT-PCR assay targeting the RBM coding sequence from 10^5^ (blue curve), 10^4^ (red curve), 10^3^ (green curve), 10^2^ (yellow curve) and 10^1^ (pink curve) RNA copy/µL. The linear regression (dashed line) in Panel A has a correlation coefficient R^2^=0.9986. C: End-point analysis of PCR fragments on gel electrophoresis. D: Electropherograms of the digestion products. Values above the peaks correspond to the size of the fragment. Peaks on the right (330bp) correspond to the homoduplexes created during the hybridization.

### Detection and identification of mutations revealing in the RBM

#### Detection of mutations in reference material derived from SARS-CoV-2 isolates

To establish the proof of concept, first experiments were conducted using viral RNA derived from cell cultures infected by three well-characterized variants: SARS-CoV-2 BavPat1, 501Y.V1 and 501Y.V2; (i) 501Y.V1 has N501Y (nt A1501T) mutation, (ii) and 501Y.V2 has E484K and N501Y (nt G1450A and A1501T, respectively) mutations by reference to the BavPat1 respectively. The theoretical cleavage profiles for pairwise combinations of the three variants are presented in Table 1. In practice PCR products were mixed in pairs in approximate equimolar quantities before denaturation/hybridization prior to the mismatch-specific endonuclease assay. The results are presented in Figure 2D. The electropherogram (blue curve) corresponding to the hybridization of the BavPat1 amplicon with itself, with no mismatch expected, resulted in a unique 330-nt long fragment. When mixed with the one of 501Y.V1 (Fig 2D, orange curve), three DNA fragments were observed at 330, 232 and 93 bp, close to the anticipated profile (Table 1). For the identification of the 501Y.V2 (grey curve), five DNA fragments can be detected, corresponding to the products from both complete and partial cleavages of the mismatches at the expected positions. Finally, when mixing amplicons from the 501Y.V1 and 501Y.V2 samples (yellow curve), three DNA fragments were observed, where the two smaller being indicative of the difference at position 484 in the Spike protein sequence of the two variants.

**Table 1:** Theoretical cleavage profiles for pairwise combinations of amplicons from BavPat1, 501Y.1 and 501Y.V2 variants. The amino-acid residue and corresponding position in the sequence of the Spike protein for each variant are in italic. Values in the table correspond the expected size of the fragments after treatment with the mismatch-specific nuclease. Underlined: fragments resulting from the complete cleavage of the heteroduplexes; Bold: fragment resulting from an incomplete cleavage.

|  | *E484 ; N501*<br>**BavPat1** | *E484 ; Y501*<br>**501Y.V1** | *K484 ; Y501*<br>**501Y.V2** |
| --- | --- | --- | --- |
| *E484 ; N501*<br>**BavPat1** | 315 nt | <u>84</u> / <u>231</u> / 315 nt | <u>51</u> / <u>84</u> / **135** / <u>180</u> / **231** / 315 nt |
| *E484 ; Y501*<br>**501Y.V1** |  | 315 nt | <u>135</u> / <u>180</u> / 315 nt |
| *K484 ; Y501*<br>**501Y.V2** |  |  | 315 nt |

#### Application to clinical samples containing SARS-CoV-2 RNA as diagnosed by routine real-time RT-PCR assay

To further assess the mutation detection method, 8 samples were tested after they were diagnosed as SARS-CoV-2 RNA positive using the routine diagnostic assay TaqPath real-time RT-PCR™ (ThermoFisher), known to discriminate the 501Y.V1 variant on the deletion observed in the S coding sequence, leading to the amplification of only two targets out the three of the test [18]. In order to make the analysis easy and rapid, it is advocated to test each sample against itself and against the selected reference with no prior quantification. In our case, the reference was the European lineage of SARS-CoV-2 (BavPat1 strain) since it was the dominant one at the time of the study. Clinical samples were identified as either SARS-CoV-2 positive for the three targets of the TaqPath assay™ (Figure 3, samples 1, 3, 4, 5, 7 and 8) or SARS-CoV-2 501Y.V1 positive (Figure 3 samples 2 and 6) with Ct ranging from 18 to 34. For samples 5 and 8, no fragment corresponding to the expected amplicon was visible in the self-hybridization assay (Figure 3, lanes 13 and 22), in line with the high Ct values obtained from initial real time RT-PCR assays. By contrast, the amplification of the target region for the other sample was satisfactory (Figure 3, lanes 1, 4, 7, 10, 16 and 19). Overall, SARS-CoV-2 positive and 501Y.V1-putative samples (2 and 6) showed profiles matching with the expectations, *ie* one mismatch with BavPat1 reference amplicon and no mismatch with 501Y.V1 reference amplicon (Figure 3, lanes 5, 6 and 17, 18 respectively).

**Figure 3:**
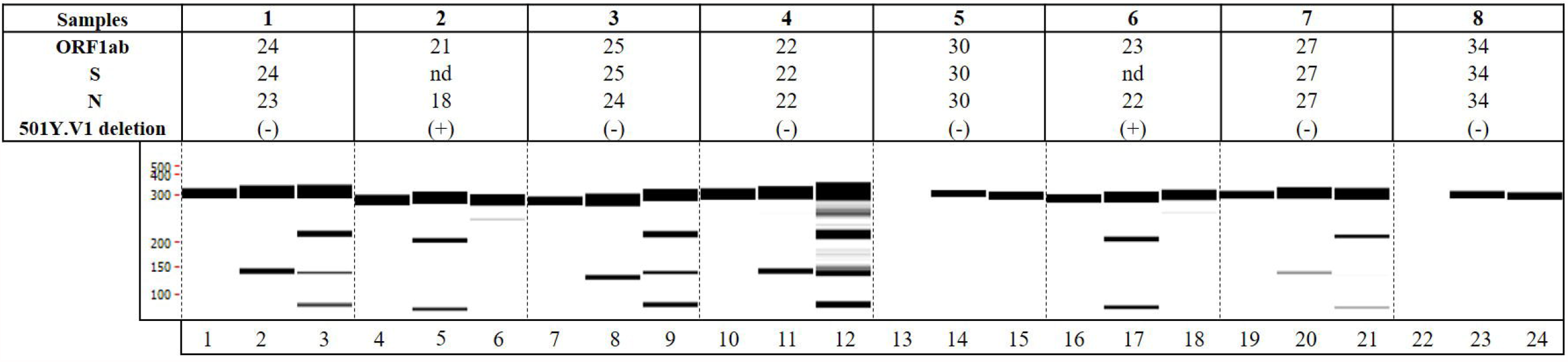
Validation of the assay on SARS-CoV-2 positive clinical samples. Ct obtained from TaqPath COVID-19 kit™ (ThermoFisher) real time RT-PCR are presented in the upper table (nd: not determined, Ct>35), leading to the following classification: (+) refers to the presence of the deletion indicative of a 501Y.V1 variant, and (-) to a non-501Y.V1 sample. Results of the mismatch-specific assay followed by capillary electrophoresis are presented under the table in a “gel-like” format. For each sample, the first lane is the auto-control (sample/sample), the second one is sample/BavPat1 and the third sample/501Y.V1.

For the other samples (Figure 3, samples 1, 3, 4 and 7), described as non-501Y.V1 according to the TaqPath assay, no cleavage was expected when compared to the BavPat1 reference amplicon, and two cleaved products with the 501Y.V1. However, at least one additional fragment was observed against both reference samples, of about 135 bp length (Figure 3, lanes 2,3,8,9,11,12,20 and 21). The PCR products were submitted to Sanger sequencing and a non-synonymous mutation yielding S477N substitution was detected, in agreement with the size of the cleaved products. This mutation had already been reported in viral populations circulating in Europe (https://www.gisaid.org/), and the corresponding variant has been shown to slightly increase the receptor binding domain’s affinity for hACE2 [19].

We next blindly evaluated the assay on 92 SARS-CoV-2 positive samples, for which Ct values were below 28 and NGS data available. The sensitivity, defined as the ability to generate an amplicon with yields compatible for the nuclease assay, is 97.83% as 2 samples out of 92 were not properly amplified. The specificity was defined as the ability to detect in the RT-PCR-positive samples a mutation compared to the BavPat1 strain or detect the lack of mutation for sequences identical to the reference. Compared to the sequence data, 2 samples out of 90 were inadequately identified, with selectivity of 97.78%. Out of the 88 remaining profiles, 82 had the same profile as in Figure 2D. Yet unmet patterns were observed and confirmed by the sequence analysis with E471D mutation detected in two samples, E484K in one sample, L452R in one sample and both L452R / N501Y in 2 others.

Altogether, these results demonstrate that the mismatch-specific nuclease assay coupled to capillary electrophoresis is a suitable assay for the detection in clinical samples of already reported mutations. The identification of atypical profiles, confirmed in this study by sequencing, also demonstrates that it is relevant for the discovery of yet unmet variants. With the incremental characterization of variants, the set of reference cleavage patterns can be updated to adapt the assay to circulating strains. As this technique can be dimensioned for 96/384 well devices and requires less than 4 hours from the extracted RNA to production of individual results, it can be used to filter clinical samples and identify those for which virus isolation and complete genome sequencing is justified for surveillance purpose.

## Conclusion

We developed a molecular assay dedicated to the surveillance of SARS-CoV-2 variants, specifically targeting the RBM coding sequence known to be involved in the functional adaptation of the virus. The assay is suitable to screen biological samples and identify the presence new or emerging mutations. The technique, based on RT-PCR amplification of the RBM coding sequence followed by mismatch-specific nuclease assay and detection by DNA capillary electrophoresis is made possible for samples with RNA titers yielding Ct values better than 28 to produce enough amplified DNA material. The procedure is amenable to high throughput and could meet the demand for large scale viral population analysis, as well as individual cases such as the surveillance of virus evolution during a chronic infection by SARS-CoV-2, a situation favourable for virus adaptation [20].

## Data Availability

no additional data

## Acknowledgements

We thank Professor Bruno Lina and the Centre National de Référence des virus des infections respiratoires for sharing extracted RNA of clinical samples. This study was funded by (i) the “European Virus Archive Global” (EVA-GLOBAL) project H2020-INFRAIA-2019 program, Project No 871029, (ii) the “Advanced Nanosensing platforms for Point of care global diagnostics and surveillance” (CONVAT), H2020, Project No 101003544. Eva Lopez is the recipient a DGA fellowship (Direction Générale de l’Armement).

## Notes

### Competing Interest Statement

The authors have declared no competing interest.

### Funding Statement

The work was supported by (i) the European Virus Archive Global (EVA-GLOBAL) project H2020-INFRAIA-2019 program, Project No 871029, (ii) the Advanced Nanosensing platforms for Point of care global diagnostics and surveillance (CONVAT), H2020, Project No 101003544

### Author Declarations

This study falls within the scope of the French Reference Methodology MR004 according to 2016.41 law dated 26 January 2016 on the modernization of the French health system. The ethics committee of University of Corsica Pascal Paoli (IRB UCPP 2020.01) approved this study

